# Meal Timing Patterns and Associations with Fat Mass in Adolescents

**DOI:** 10.64898/2026.04.22.26351498

**Authors:** Jessica E. Decker, Knashawn H. Morales, Pin-Wei Chen, Lindsay Master, Misol Kwon, Erica C. Jansen, Babette S. Zemel, Jonathan A. Mitchell

## Abstract

**Background:** The timing of energy intake could be important in the development of obesity. However, most observational evidence stems from adults, anthropometric defined obesity outcomes, single meal timing phenotyping, and traditional regression modeling.

**Objective:** We aimed to describe meal timing patterns in adolescents and determine if they associated with fat mass by modeling the median and all other percentiles of the frequency distribution.

**Methods:** We analyzed data from the Sleep and Growth Study 2 (S-Grow2, N=286, 12-13y). Participants completed 3-day 24-hour dietary recalls and time stamped eating occasions were used to define 8 meal timing traits, with aide from self-reported wake and bed timing. Principal component analysis (PCA) identified multi-dimensional meal timing patterns. Fat mass index (FMI) was estimated using dual energy X-ray absorptiometry. Quantile regression assessed if there were associations between meal timing traits and FMI across the entire FMI frequency distribution.

**Results:** The typical first and last eating occasions were 8:00am (40 minutes after waking) and 8:00pm (2.7 hours before sleep), respectively, thus the eating period typically lasted 11.5 hours per day. The typical eating period midpoint was 2:15pm, and the timing when 50% of energy intake was consumed typically occurred at 3:15pm. PCA revealed three meal timing patterns: 1) “Delayed Start, Condensed Eating Period” (43% of variance; shorter eating period and delayed timing of first eating); 2) “Late, Sleep Proximal Eating” (30% of variance; later timing of last eating and extended eating period), and 3) “Later Energy Intake” (10% of variance; delayed energy intake midpoint). Higher scores for the “Delayed Start, Condensed Eating Period” pattern associated with higher body mass index and FMI at the upper tails of their distributions.

**Conclusions:** Distinct multidimensional meal timing patterns emerged in early adolescence, with the delayed start, condensed eating period pattern potentially associated with higher adiposity.

**Clinical Trials Registry Site and Number:** N/A

## Introduction

Obesity is a leading public health problem, linked to multiple chronic diseases that can originate in childhood^1-4^. An estimated 17% of children have obesity and the prevalence is especially high among adolescents (22%)^5^. To help manage and prevent childhood obesity, the Dietary Guidelines for Americans (DGAs) recommends increasing the consumption of nutrient dense foods (e.g., fruits, vegetables, whole grains, lean proteins) and limiting consumption of energy dense foods (e.g., foods high in added sugars, sodium, and saturated fats)^1-4^. Notably, the current dietary guidelines do not include information pertaining to the timing of energy intake. This may be an important omission since emerging evidence suggests that when food is consumed within each 24-hour daily cycle can be obesogenic^6-12^.

Compelling evidence linking the timing of energy intake to obesity and related metabolism stems from time restricted feeding experiments involving animal models^13, 14^ and adults^7, 15, 16^. For example, mice fed a high fat diet ad-libitum gain excess fat mass; however, when the same high fat diet was restricted to an 8–10-hour nocturnal feeding window the excess weight gain and impaired metabolic consequences were prevented^13^. In adults, experiments restricting the feeding period to 9 or 10 hours per day have revealed improvements in obesity-related outcomes, such as weight and body mass index (BMI)^8, 10^. Such restriction can prevent energy intake late at night, which is postulated to be obesogenic due to circadian misalignment. However, experiments in adults involving night shift workers have also shown metabolic health benefits with time restricted feeding, suggesting restricted feeding benefits regardless of the time of day or night^16^.

Given this evidence, there is a need to determine if meal timing patterns are contributing to childhood obesity, especially in adolescence when pediatric obesity rates peak^5^, pubertal onset and progression is occurring with phase shift delays in sleep-wake patterns^17^, and when children become more independent with respect to what and when they eat^18^. Before designing experimental studies, we first need to observe adolescents to assess variability in their meal timing patterns and test if there are associations with obesity-related outcomes. Three prior observational studies involving adolescents have been limited to anthropometric defined obesity outcomes, single meal timing phenotyping, and traditional regression modeling^19-21^. Extending this area of research, we leveraged dual energy X-ray (DXA) estimated fat mass data, multi-dimensional meal timing phenotyping, and quantile regression modeling. We aimed to describe multi-dimensional meal timing patterns in adolescents and determined if meal timing patterns associated with fat mass outcomes by modeling entire frequency distributions for each fat mass outcome variable.

## Methods

### Sample

Cross-sectional data from the Sleep and Growth Study 2 (S-Grow2) were analyzed. This study enrolled typically developing adolescents without a known medical (e.g., cancer, gastroenterology, and sleep disorders) and/or behavioral (e.g., anxiety, depression, and ADHD) condition from the southeastern Pennsylvania and southern New Jersey regions from 2021-2024. All participants were in 7^th^ grade (12-13 years old). Written informed consent and assent were obtained from parents or guardians and adolescents, respectively, and the study protocol was approved by the Institutional Review Board at the Children’s Hospital of Philadelphia (CHOP; IRB: 20-17328).

### Obesity-Related Outcomes

Each participant visited the Children’s Hospital of Philadelphia’s Nutrition Core Lab, where trained staff used a wall-mounted stadiometer to measure height (to the nearest 0.1cm) and a digital scale to measure weight (to the nearest 0.1 kg) in triplicate. Body mass index (BMI, kg/m^2^) was calculated for each participant, and BMI defined overweight and obesity were used for descriptive purposes using age- and sex-specific International Obesity Task Force standards. Participants completed a whole-body DXA scan (Hologic Discovery A; Hologic Inc.; Bedford, MA), to estimate total fat mass (kg) and allow for the calculation of fat mass index (FMI, kg/m^2^).

### Dietary Recall Methods

Following the study visit, participants entered a 2-week observational period at home. During this period, registered dietitians called adolescents to capture up to three days of 24-hour dietary recalls. The multiple-pass method was used to guide participants to recall the timing and amounts (in household measures) of foods and beverages consumed from midnight-to-midnight during the prior day. These data were entered into the Nutrition Data System for Research database (version 2024) and converted into grams of foods and beverages consumed. Of the output files generated, the “Meal File” provided time stamped meal data, the “Daily Intake Totals File” provided estimates of energy (kcal/day) and macronutrient (grams/day) intake, and the “HEI-2015 Daily Intake Totals File” provided Healthy Eating Index (HEI) 2015 scores (HEI-2015; range: 0-100, with higher scores indicating greater diet quality)^22^.

We removed days that were noted as unreliable from the dietitian at the end of the phone call (e.g., if a child was ill and reported that the recall day was not a typical day). Due to the midnight-to-midnight (and not wake-to-wake) reporting structure of the 24-hour dietary recall, we identified eating occasions reported between midnight and 5:00am to check if these were first or last meals by cross-checking with sleep diary time in bed data; we recoded these as last meals when the meal was consumed prior to bedtime.

Using the cleaned meal timing data, the following variables were calculated: 1) time between waking and first eating occasion (hours), 2) time of the first eating occasion (clock time), 3) time of eating period midpoint (clock time), 4) time of 50% daily energy intake (clock time), 5) time of last eating occasion (clock time), 6) time between last eating occasion and sleep onset (hours), 7) number of eating occasions, and 8) total time of the eating period (hours). The waking and bedtime data were captured from self-reported sleep diaries.

### Covariates

Age was included as a covariate since older age has been associated with later meal timing^23^ and an increased prevalence of obesity^24^. Sex differences in meal timing have been documented^25^, and in regard to the prevalence of obesity and so was included as a covariate. HEI and energy intake (estimated from 3-day 24-hour diet recalls) were included as covariates since lower diet quality and excess energy intake have been associated with meal timing and a greater prevalence of obesity^26^. Physical inactivity and shorter sleep duration may associate with meal timing and are known to associate with obesity^19, 20^; self-report physical activity and sleep duration were included as covariates. Finally, social jet lag was included as a covariate as a marker of circadian misalignment since this may associate with meal timing and obesity; participants wore a GENEActiv actigraphy device for up to 2-weeks and the data were processed using the GGIR R-package to estimate sleep timing^27^. Social jet lag was calculated as the difference between mid-sleep point on weekend days and weekdays, corrected for potential sleep debt.

### Statistical Analysis

Descriptive statistics (frequencies and percentages for categorical variables; mean and standard deviation for continuous variables) were used to summarize the data. Principal component analysis (PCA) was performed on the eight meal timing traits to identify multi-dimensional meal timing patterns. Each individual meal timing trait was averaged across the total number of recall days available and standardized within the sample. An eigen value of 1.0 and visual inspection of the scree plot were used to determine the maximum number of principal components to retain. The standardized loading scores for each principal component were used as the primary exposure variables. Pairwise correlation was used to determine if PCA derived meal timing patterns correlated with energy intake, macronutrient intake, and HEI-2015 scores.

Quantile regression was used to determine if the PCA-derived meal timing patterns associated with FMI and BMI. Instead of modeling the mean, this statistical approach models the median and any other percentile across the frequency distribution. Quantile regression is therefore well suited for modeling outcomes that do not follow a normal distribution as the percentile specific associations obtained are not impacted by outliers. Further, this method allows for the detection of differential associations across the frequency distribution without relying on categorization.

This is important because, in the context of obesity, associations detected at the upper percentiles of FMI and BMI frequency distributions are of relevance. Models were performed that were minimally adjusted (model 1 was adjusted for age and sex) and fully adjusted (model 2 was additionally adjusted for diet quality, total energy intake, physical activity, sleep duration, and social jet lag).

In addition, we completed two sensitivity analyses to examine whether associations between meal timing and the obesity outcomes differed by sex and degree of social jetlag. This was accomplished by repeating the quantile regression models by each sex and using the median split for social jetlag. Stata 17.0 (StataCorp, College Station, TX) was used for all analyses except for the quantile regressions, for which Stata 19.0 was used.

## Results

A total of 321 adolescents completed a DXA scan and started the home observational period. Five participants did not provide any 24-hour diet recall data, 3 participants did not contribute any reliable recall days, 18 participants did not complete their 24-hour recalls during the same period as which the sleep diaries were collected, and 9 participants did not complete any weekend sleep diaries. These participants were removed, yielding a final analytical sample of 286 adolescents (**Figure 1**) contributing 714 days of recall and 3,000 eating occasions. In the analytical sample, 4 eating occasions occurred between midnight and 5:00 am that were initially classed as first meals; after reviewing the sleep diary bedtime data these eating occasions were confirmed to have taken place before going to bed and so were recoded as last meals.

**Figure 1.**
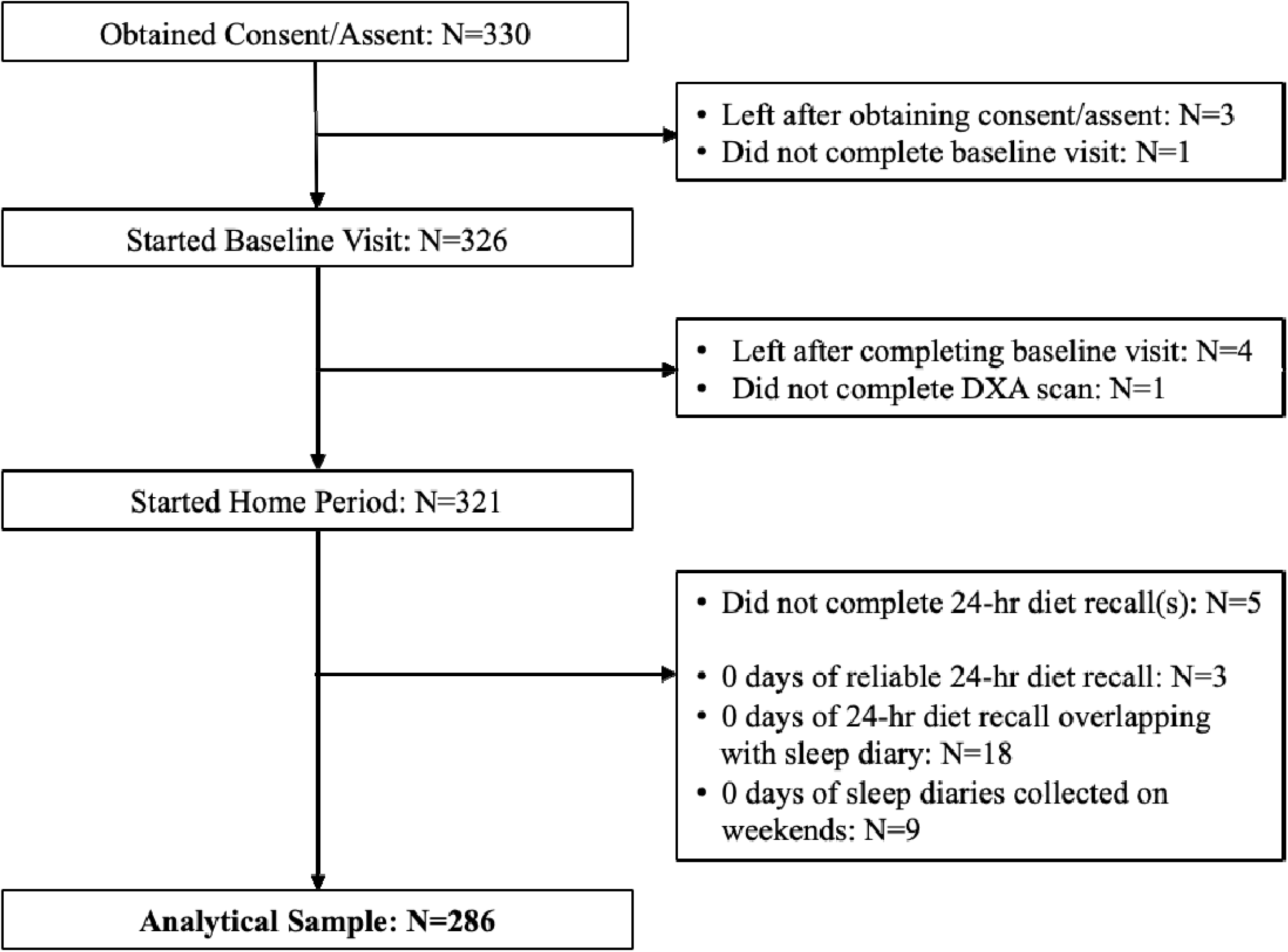
Flow chart of participants included in analysis.

The mean age of the analytical sample was 12 years, 54% were male, 66% identified as White, and 35% of adolescents resided in a home with an annual household income between $100,000-$199,999 (Table 1). The proportion of adolescents with BMI defined obesity was 12% and the mean HEI score was 47 (Table 1).

**Table 1.** Baseline characteristics of the sample.

|  | **Overall** |  | **Female** | **Male** |  | **SJL < 50<sup>th</sup>** | **SJL ≥ 50<sup>th</sup>** |
| --- | --- | --- | --- | --- | --- | --- | --- |
| N | 286 |  | 131 | 155 |  | 143 | 143 |
| Age, years, mean (SD) | 12.4 (0.5) |  | 12.4 (0.5) | 12.4 (0.5) |  | 12.4 (0.5) | 12.4 (0.5) |
| Female, N (%) | 131 (45.8) |  | 131 (100) | 0 (0) |  | 62 (43.4) | 69 (48.3) |
| Male, N (%) | 155 (54.2) |  | 0 (0) | 155 (100) |  | 81 (56.6) | 74 (51.7) |
| Asian, N (%) | 13 (4.5) |  | 3 (2.3) | 10 (6.5) |  | 11 (7.7) | 2 (1.4) |
| Black, N (%) | 52 (18.2) |  | 21 (16.0) | 31 (20.0) |  | 20 (14.0) | 32 (22.4) |
| White, N (%) | 188 (65.7) |  | 92 (70.2) | 96 (61.9) |  | 97 (67.8) | 91 (63.6) |
| 2+ Races, N (%) | 26 (9.1) |  | 10 (7.6) | 16 (10.3) |  | 12 (8.4) | 14 (9.8) |
| Other, N (%) | 3 (1.0) |  | 1 (0.8) | 2 (1.3) |  | 2 (1.4) | 1 (0.7) |
| Unknown, N (%) | 4 (1.4) |  | 4 (3.1) | 0 (0.0) |  | 1 (0.7) | 3 (2.1) |
| Hispanic, N (%) | 260 (90.9) |  | 119 (90.8) | 141 (91.0) |  | 129 (90.2) | 131 (91.6) |
| Non-Hispanic, N (%) | 13 (4.5) |  | 7 (5.3) | 6 (3.9) |  | 7 (4.9) | 6 (4.2) |
| Unknown, N (%) | 13 (4.5) |  | 5 (3.8) | 8 (5.2) |  | 7 (4.9) | 6 (4.2) |
| Household Income |  |  |  |  |  |  |  |
| <\$50,000, N (%) | 25 (8.7) | | 12 (9.2) | 13 (8.4) | | 12 (8.4) | 13 (9.1) |
| \$50,000-\$99,999, N (%) | 41 (14.3) | | 17 (13.0) | 24 (15.5) | | 14 (9.8) | 27 (18.9) |
| \$100,000-\$199,999, N (%) | 100 (35.0) | | 50 (38.2) | 50 (32.3) | | 54 (37.8) | 46 (32.2) |
| At least \$200,000, N (%) | 108 (37.8) | | 46 (35.1) | 62 (40.0) | | 57 (39.9) | 51 (35.7) |
| Unknown, N (%) | 12 (4.2) |  | 6 (4.6) | 6 (3.9) |  | 6 (4.2) | 6 (4.2) |
| Body Mass Index Category |  |  |  |  |  |  |  |
| Underweight, N (%) | 16 (5.6) |  | 6 (4.6) | 10 (6.5) |  | 7 (4.9) | 9 (6.3) |
| Normal Weight, N (%) | 183 (64.0) |  | 86 (65.6) | 97 (62.6) |  | 90 (62.9) | 93 (65.0) |
| Overweight, N (%) | 53 (18.5) |  | 24 (18.3) | 29 (18.7) |  | 29 (20.3) | 24 (16.8) |
| Obese, N (%) | 34 (11.9) |  | 15 (11.5) | 19 (12.3) |  | 17 (11.9) | 17 (11.9) |
| Fat Mass Index, kg/m<sup>2</sup>, mean (SD) | 5.0 (2.9) |  | 5.6 (2.7) | 4.6 (3.0) |  | 5.1 (3.0) | 5.0 (2.8) |
| Healthy Eating Index, mean (SD) | 46.7 (13.8) |  | 45.4 (13.3) | 47.8 (14.2) |  | 46.0 (14.4) | 47.4 (13.1) |
| Energy Intake, kcal/day, mean (SD) | 1836.3 (592.3) |  | 1738.4 (482.9) | 1919.1 (661.4) |  | 1881.7 (631.0) | 1791.0 (549.4) |
| Physical Activity, hours/day, mean (SD) | 2.0 (1.4) |  | 1.9 (1.3) | 2.2 (1.5) |  | 2.2 (1.5) | 1.9 (1.3) |
| Total Sleep Time, hours/night, mean (SD) | 8.4 (1.3) |  | 8.2 (1.3) | 8.4 (1.2) |  | 8.2 (1.3) | 8.5 (1.2) |
| Social Jet Lag < 50<sup>th</sup> Percentile, N (%) | 143 (50.0) |  | 62 (47.3) | 81 (52.3) |  | 143 (100) | 0 (0) |
| Social Jet Lag ≥ 50<sup>th</sup> Percentile, N (%) | 143 (50.0) |  | 69 (52.7) | 74 (47.7) |  | 0 (0) | 143 (100) |

Abbreviations: SJL, social jet lag.

The median timing of the first eating occasion was 08:00 (range: 05:00-16:30; SD: 1.9 hours), 0.66 hours (SD: 1.9 hours) after waking (Figure 2). The median timing of the last eating occasion was 20:00 (range: 14:00-00:00; SD: 1.4 hours), 2.7 (SD: 1.8) hours before sleep (Figure 2). The median duration of the eating period was 11.5 (range: 2.5-16.5; SD: 2.5) hours per day, and the median eating period midpoint occurred at 14:00 (range: 11:47-18:20; SD: 1.2 hours) (Figure 2). The median timing of when half of the days’ total energy intake was consumed was 14:50 (range: 08:00-23:50; SD: 2.8 hours) (Figure 2).

**Figure 2.**
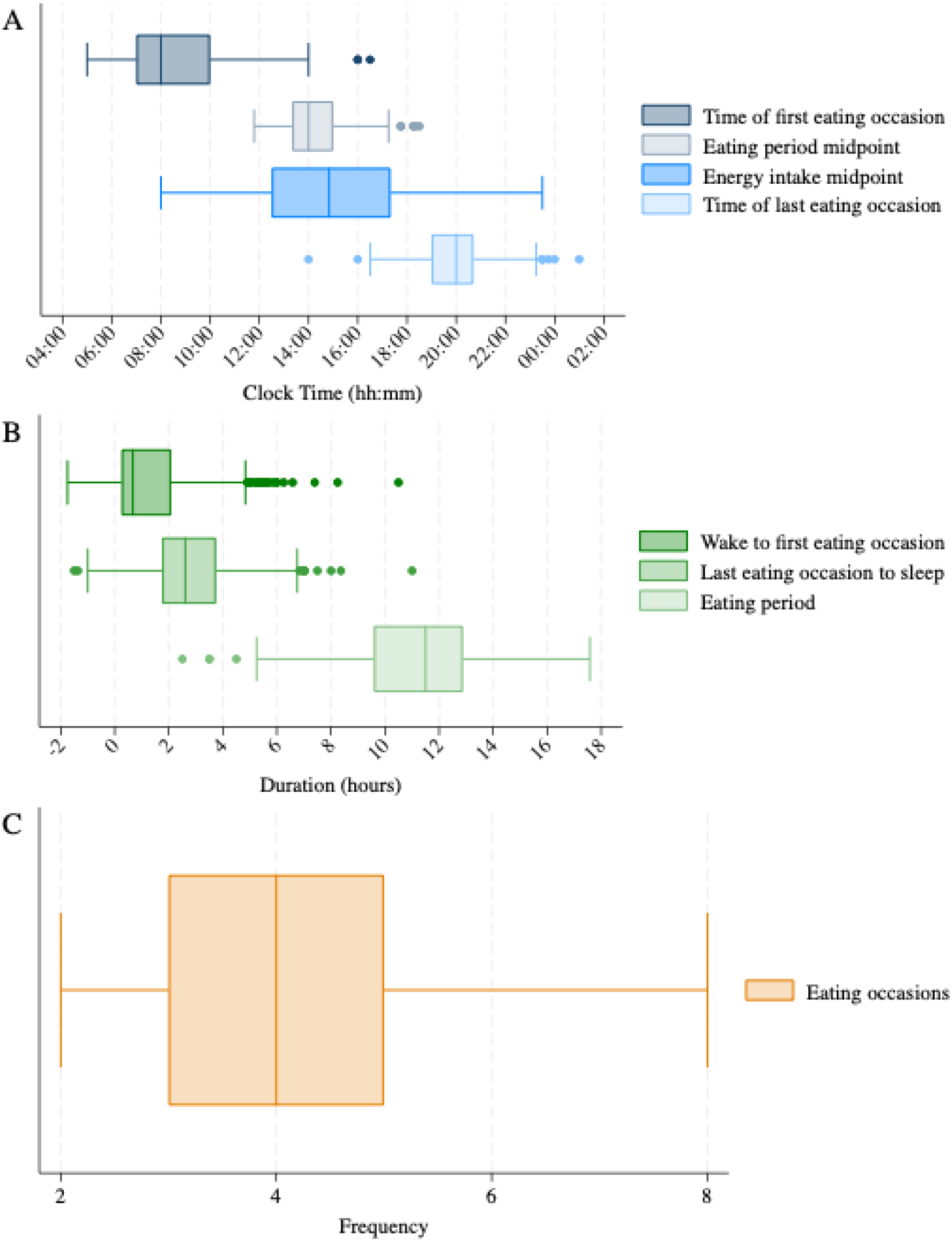
Descriptive statistics for the individual meal timing traits.

The PCA revealed three distinct meal timing patterns that accounted for 83% of the variation in meal timing (**Supplemental Figure 1**). Individuals with higher PC1 loading scores were characterized by a condensed eating period, later morning eating onset, and lower number of eating occasions (**Table 2**). We therefore refer to PC1 as the “Delayed Start, Condensed Eating Period” pattern. A higher score for this pattern was negatively correlated with HEI-2015 scores (r=-0.18), energy intake (r=-0.15), and carbohydrate intake (r=-0.23), and positively correlated with protein intake (r=0.14) (**Supplemental Figure 2**). Individuals with higher PC2 loading scores were characterized by an extended eating period and a later timing of eating within close proximity to sleep, and more eating occasions (**Table 2**). We therefore refer to PC2 as the “Late, Sleep Proximal Eating” pattern. A higher score for this pattern was positively correlated with energy intake (r=0.19), carbohydrate intake (r=0.18), and fat intake (r=0.17) (**Supplemental Figure 2**). Individuals with higher PC3 loading scores were characterized by an extended eating period with most calories consumed in the later afternoon and evening (**Table 2**). We therefore refer to PC3 as the “Later Energy Intake” pattern. This meal timing pattern was not correlated with HEI-2015 score, energy intake, or macronutrient intake (**Supplemental Figure 2**).

**Table 2.** Factor loadings for each retained principal component (PC).

|  | **PC1** | **PC2** | **PC3** |
| --- | --- | --- | --- |
| Narrative name | “Delayed Start,<br>Condensed Eating<br>Period” | “Late, Sleep<br>Proximal Eating” | “Later Energy<br>Intake” |
| Variance explained | 43.4% | 29.8% | 9.7% |
| Factor Loadings |  |  |  |
| Wake to first eating occasion | 0.43 | 0.12 | -0.12 |
| Time of first eating occasion | 0.50 | 0.15 | -0.13 |
| Eating period midpoint | 0.34 | 0.46 | -0.19 |
| Energy intake midpoint | 0.19 | 0.28 | 0.93 |
| Time of last eating occasion | -0.14 | 0.59 | -0.15 |
| Last eating occasion to sleep | 0.22 | -0.50 | 0.15 |
| Number of eating occasions | -0.32 | 0.16 | 0.11 |
| Eating period | -0.49 | 0.21 | 0.02 |

For PC1 (“Delayed Start, Condensed Eating Period”), no associations with FMI or BMI were detected in the lower half of their frequency distributions (Figure 3, fully adjusted; and Supplementary Figure 3, minimally adjusted). However, at the 60^th^ percentile and above for FMI and BMI, the point estimates for a higher PC1 loading score steadily increased with associations detected at the 94^th^ to 99^th^ percentiles (e.g., 1 SD increase in PC1 loading score in the fully adjusted model associated with 2.03 higher FMI at the 99^th^ percentile; 95% CI: 0.09, 4.0). These patterns across the FMI and BMI percentiles were generally consistent for male and female adolescents (Supplementary Figures 4 and 5), but more prominent for adolescents with higher social jet lag (Supplementary Figures 6 and 7).

**Figure 3.**
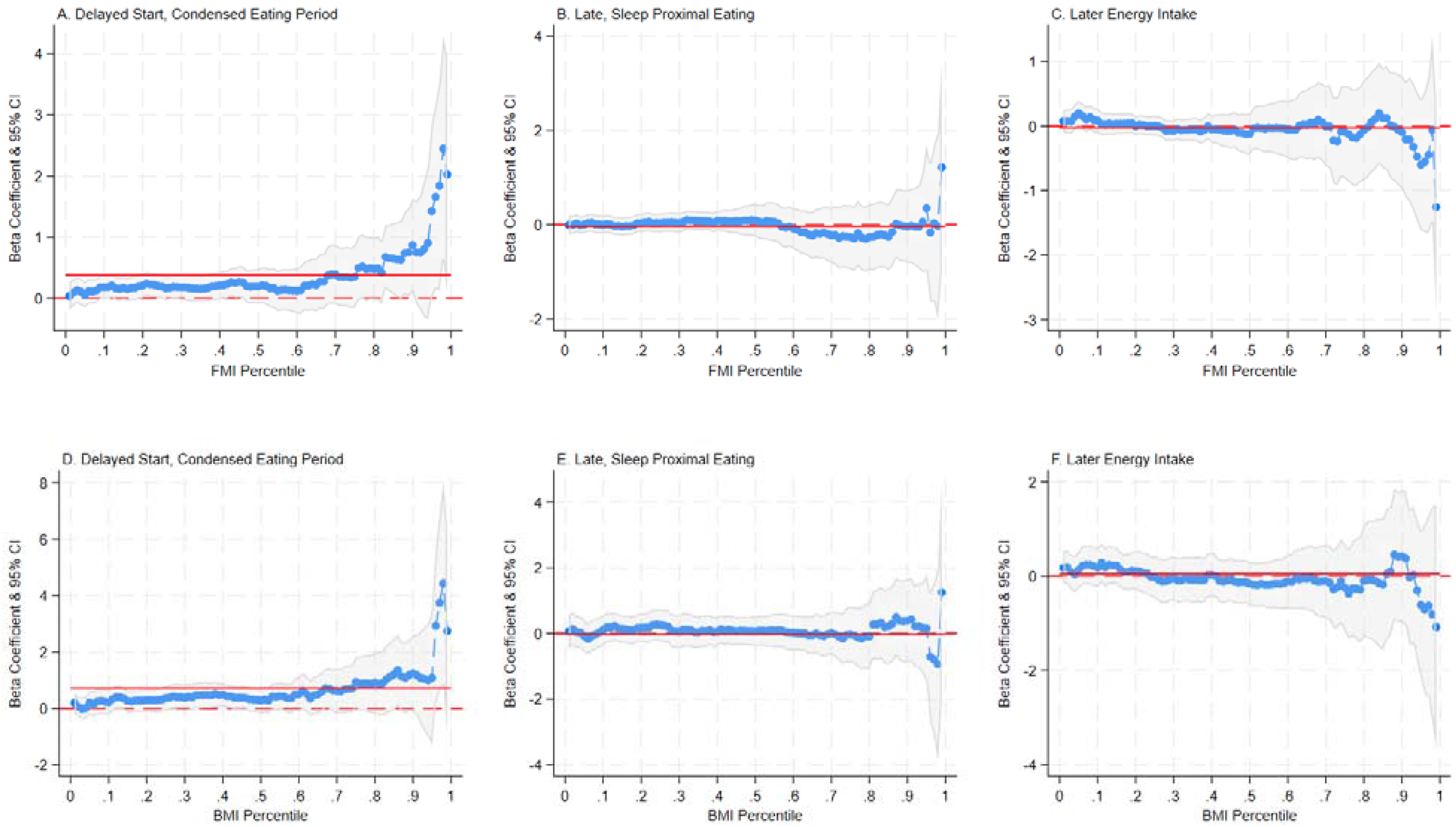
Quantile regression plots illustrating fully adjusted associations (beta, 95% confidence interval) between three meal timing patterns and obesity outcome metrics. All models were adjusted for age, sex, diet quality, energy intake, physical activity, sleep, and social jet lag. The red solid line represents the ordinary least squares estimate. The red dotted line indicates the reference line. Corresponding minimally adjusted data are given in Supplementary Figure 3. Abbreviations: BMI, body mass index; FMI; fat mass index.

For PC2 (“Late, Sleep Proximal Eating”), no associations with FMI or BMI were detected in the lower or upper half of their frequency distributions (Figure 3, fully adjusted; and Supplementary Figure 3, minimally adjusted). This remained for both social jetlag groups (Supplementary Figures 6 and 7). However, sex differences were detected (Supplementary Figures 4 and 5). For male adolescents at the 60^th^ percentile and above for FMI and BMI, the point estimates for a higher PC1 loading score steadily decreased with associations detect at the 94^th^ to 99^th^ percentiles (e.g., 1 SD increase in PC2 loading score associated with 2.4 lower FMI at the 99^th^ percentile; 95% CI: -3.69, -1.17).

For PC3 (“Later Energy Intake”), no associations with FMI or BMI were detected in the lower or upper half of their frequency distributions (Figure 3, fully adjusted; and Supplementary Figure 3, minimally adjusted). This remained for both sexes (Supplementary Figures 4 and 5) and social jetlag groups (Supplementary Figures 6 and 7).

## Discussion

There is emerging evidence that the timing of when food is consumed may be important for obesity prevention and treatment^6-12^. However, this has not been extensively investigated in the context of adolescent obesity. Using time stamped meal data from 3-day 24-hour dietary recalls, we described meal timing patterns in early adolescence and used quantile regression to determine if they associated with fat mass by considering the entire frequency distributions of FMI and BMI. The “Delayed Start, Condensed Eating Period” pattern was characterized by a shorter eating period due to a later timing of eating onset; a higher score for this pattern associated with higher FMI and BMI at the upper end of the frequency distributions. The “Late, Sleep Proximal Eating” pattern was characterized by a longer eating period with later timing of eating at night in close proximity to going to bed; this pattern was not associated with FMI or BMI, although for male adolescents a higher score was associated with lower FMI and BMI at the upper tails of the frequency distributions. Finally, the “Later Energy Intake” pattern was characteristic of most energy intake occurring in the later afternoon and evening; this pattern as not associated with FMI or BMI.

We observed that adolescents aged 12-13y start eating 40 minutes after waking, around 08:00, with the eating period lasting on average for 11.5 hours. They typically stopped eating at 20:00, approximately 2.6 hours before sleep onset. This is aligned with findings from other studies. Using 3-day 24-hour diet recalls, five studies reported that children and adolescents typically begin a 10-13 hour eating period eating around 08:30 and finish around 20:00^28-33^. We also observed that the 12-to-13-year-old adolescents in our sample had energy intake and eating period midpoints that typically occurred at 14:50 and 14:00, respectively. Mota et al. used 2-day food diaries and found children and adolescents 10-19 years typically had an eating period midpoint timing of approximately 14:00, but an energy intake midpoint timing of approximately 11:30^30^. The average eating period midpoint time aligns with our data, but the energy intake midpoint time was several hours earlier. This collective evidence indicates a relatively high degree of consistency in using dietary recalls and food diaries to estimate meal timing in adolescents. However, not all studies included sleep data to derive estimates of the timing of first and last meals in proximity to sleep onset and offset. The midnight-to-midnight recall method also is not ideal for meal timing capture and a wake-to-wake model may be an alternative approach to enhance this method from a chrononutrition standpoint.

Through PCA, three multi-dimensional meal timing patterns were revealed. The first component - the “Delayed Start, Condensed Eating Period” pattern – characterized participants as having a greater tendency of having a shorter eating period, with a longer delay to eating after waking, and eating less frequently throughout the day. The second - “Late, Sleep Proximal Eating” – was a pattern characterized by a longer feeding period and a shorter time between their last meal and bedtime. The third pattern – “Later Energy Intake” – was a pattern where most energy intake occurred in the later afternoon and evening. The “Delayed Start, Condensed Eating Period” pattern was weakly and negatively correlated with energy intake and carbohydrate intake but positively correlated with protein intake. The “Late, Sleep Proximal Eating” pattern was positively correlated with energy, carbohydrate, and fat intake. Due to the sample-specific nature of the meal timing patterns we identified, comparisons to the limited existing research regarding meal timing and obesity in children are challenging. Nonetheless, others using similar techniques identified meal timing patterns. Palla et al. used principal component analysis and identified diurnal energy intake patterns characterized by greater energy intake:1) between 01:00-02:00, 14:00-15:00, and after 21:00; 2) dispersed from 05:00-20:00, and 3) greatest in the afternoon, around 13:00 and 18:00^11^. In adult participants of the National Health and Nutrition Examination Survey, Kim et al used latent class analysis to identify six chrononutrition patterns, which included patterns such as “restricted eating window”, “extended window eating”, and “later heavy eating”^33^.

This is the first study to use quantile regression to assess the relationship between meal timing patterns and DXA-derived fat mass. This is advantageous, as it allows us to assess the relationship between meal timing patterns towards the upper tail of the fat mass distribution, which is of particular interest in the context of pediatric obesity as it is unlikely that such an association is uniform across the fat mass distribution. A higher score for the “Delayed Start, Condensed Eating Period” pattern would be comparable to time restricted feeding or prolonged overnight fast, whereas a lower score for the “Late, Sleep Proximal Eating” patterns would be comparable to time restricted feeding or prolonged overnight fast. A number of studies report metabolic and weight benefits of such time restricted feeding and prolonged overnight fast in adults^9, 34-38^, although not all studies support such associations^36^. Interestingly, we observed in early adolescence associations between a higher “Delayed Start, Condensed Eating Period” score with higher FMI and BMI at the upper tails of their distributions. A prior cross-sectional study in adolescents reported null associations between the timing of the first and last eating occasions and odds of having BMI defined overweight or obesity^30^. However, others have observed associations between longer eating periods and higher BMI^30, 32^ but lower fat mass^32^. Mota et al. found that children consuming a higher percent of total calories after 21:00 were 20% more likely to have overweight or obesity^30^. Palla et al. found that a meal timing pattern characterized by early eating, an extended eating period and later energy intake was not associated with mean BMI^11^.

In a sensitivity analysis, we examined whether the relationship between meal timing patterns and fat mass was consistent between males and females. Overall, we found no sex differences between the “Delayed Start, Condensed Eating Period” and “Later Energy Intake” meal timing patterns with respect to FMI or BMI. However, we did detect a stronger association between the “Late, Sleep Proximal Eating” and FMI and BMI at the upper tails of their frequency distributions. Our participants were in early adolescence, and females are more likely to start puberty sooner than males^39^. A shift towards a later chronotype is common as adolescents reach puberty. It is possible, then, that the sex difference we identified was simply due to males not having reached puberty yet. Future studies should examine the association between meal timing patterns and fat mass between male and female sex during and after puberty has surpassed.

We also examined whether associations between meal timing patterns and obesity outcomes were consistent regardless of social jet lag category. There were no social jetlag distinct associations between any of the three meal timing patterns and the obesity outcomes. This suggests that in early adolescence, those with misaligned social and biological clocks are not more sensitive to the obesogenic effects of when meals are consumed. However, more studies are needed to investigate this relationship, especially in older adolescence when social jet lag may be more pronounced.

Our study has limitations. This was a single-site, cross-sectional study, limited to early adolescence. Future studies are needed to independently replicate and expand our findings, especially through longitudinal designs from early to late adolescence when meal timing patterns likely change during puberty in parallel to changes in sleep timing (i.e. sleep phase delay and more opportunities to eat later in the day). The proportion of our sample with BMI defined obesity was 14%, slightly below the national average for adolescence; given the associations detected at the upper tails of the FMI and BMI distributions, we may have detected more robust signals had we had a higher proportion of obesity included in the sample. The self-report approach we used to capture meal timing is reliant on recall; it would be of interest to identify alternative methods to overcome this limitation in the future; for example, continuous glucose monitoring may provide a sensor-based method to help quantify meal timing. Additionally, 24-hour diet recalls use a midnight-to-midnight assessment period and a wake-to-wake approach is more suitable for chrononutrition purposes. Finally, the standard three day approach is also limiting and acquiring additional days of data to robustly assess meal timing on weekend and weekend days is needed^40, 41^.

## Conclusion

In our sample representing early adolescents, we used 24-hour diet recalls and sleep diaries to describe meal timing and used PCA to identify three multi-dimensional meal timing patterns. At the upper tails of the FMI and BMI distributions, the “Delayed Start, Condensed Eating Period” pattern was associated with higher FMI and BMI, and limited to male adolescents, the “Late, Sleep Proximal Eating” pattern was associated with lower FMI and BMI. Meal timing could contribute to the early development of obesity and warrant further investigation as we seek strategies to prevent and treat childhood obesity.

## Abbreviations

BMI: Body Mass Index
CHOP: Children’s Hospital of Philadelphia
DGAs: Dietary Guidelines for Americans
DXA: Dual Energy X-ray Absorptiometry
FMI: Fat Mass Index
HEI: Healthy Eating Index
PCA: Principal Component Analysis
S-Grow2: Sleep and Growth Study 2
US: United States

## Acknowledgements

JD and JM designed and conducted the research and analyzed the data. JD, KM, PC, LM, MK, EJ, BZ, and JM wrote the paper. JD had primary responsibility for the final content. All authors read and approved the final manuscript.

## Data Availability

Data described in the manuscript, code book, and analytic code will be made available upon pending request approval.

## Declaration of Generative AI and AI-assisted technologies in the writing process

During the preparation of this work the authors used CHOP GPT in order to improve statement readability. After using this tool/ service, the author(s) reviewed and edited the content as needed and take full responsibility for the content of the publication.

