## Supplementary File for "Meal Timing Patterns and Associations with Fat Mass in Adolescents"

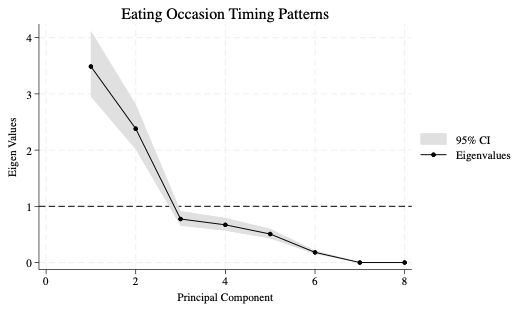

**Supplemental Figure 1.** Scree plot from the principal component analysis.

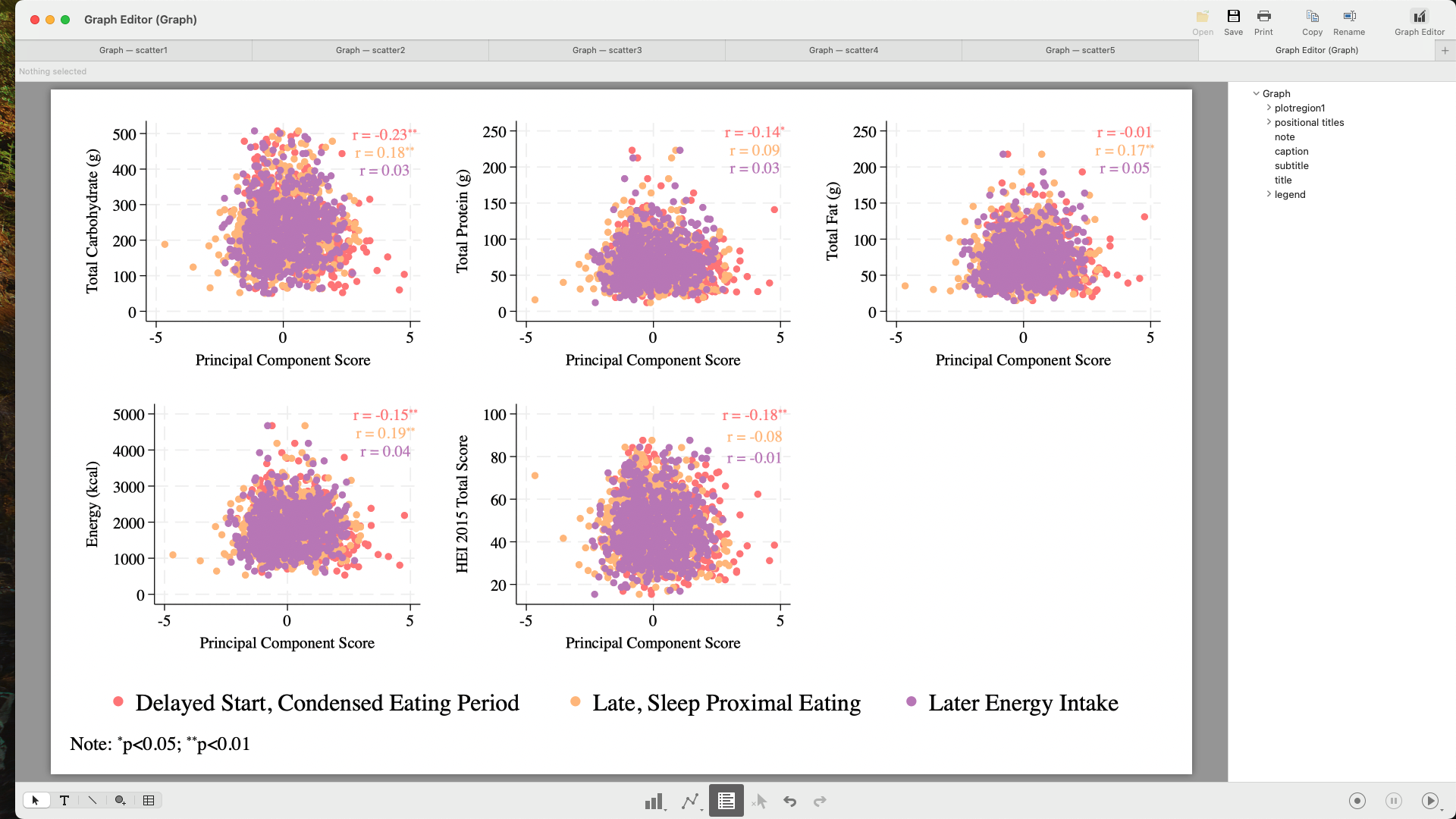

**Supplemental Figure 2.** Scatterplots and correlations between principal component loading scores with energy, macronutrients, and diet quality.

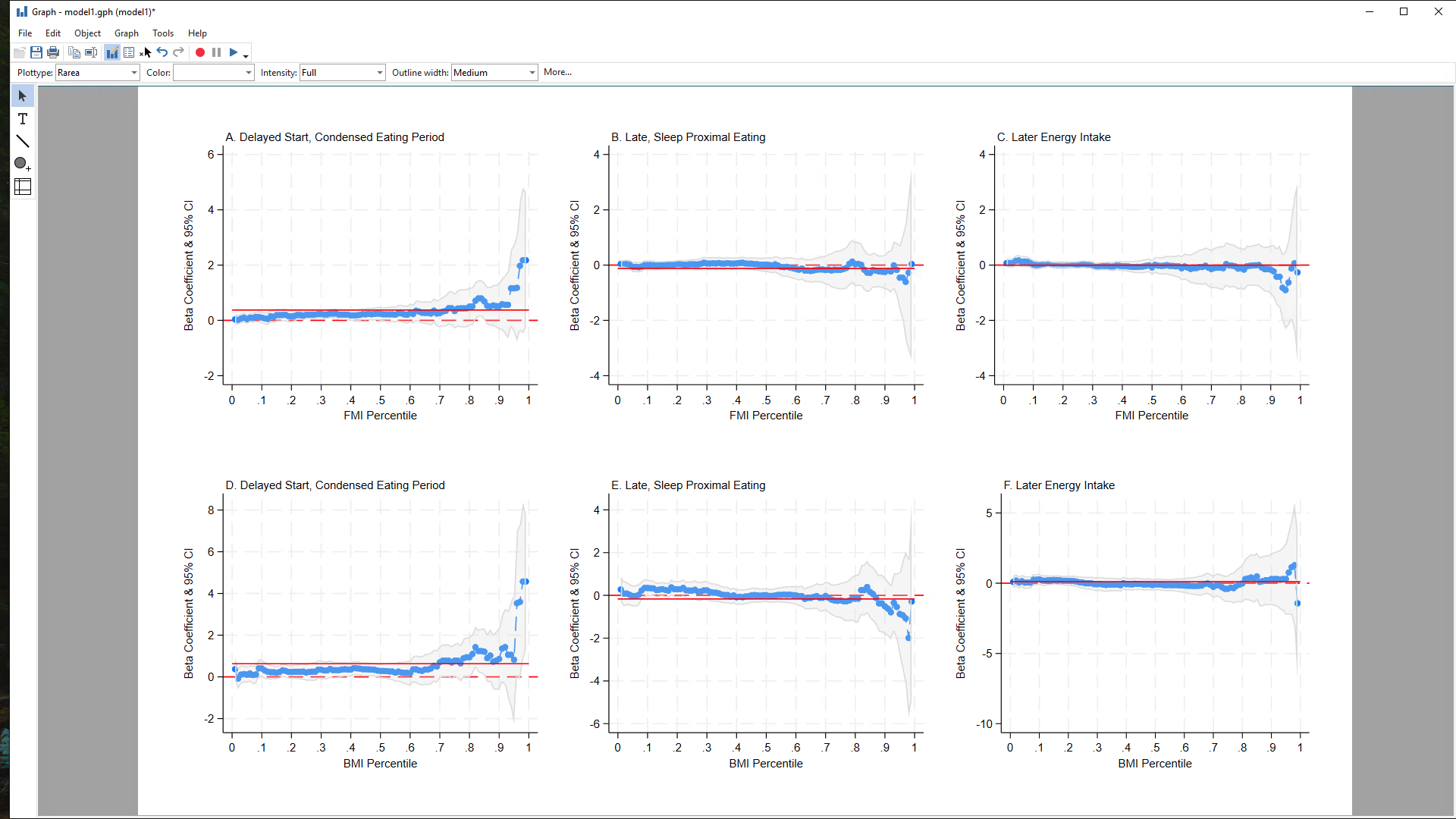

**Supplemental Figure 3.** Quantile regression plots illustrating minimally adjusted associations between three meal timing patterns obesity outcome metrics. All models were adjusted for age and sex, diet quality, energy intake, physical activity, sleep, and social jet lag. The red solid line represents the ordinary least squares estimate. The red dotted line indicates the reference line. Corresponding fully adjusted data are given in Figure 3. Abbreviations: BMI, body mass index; FMI; fat mass index.

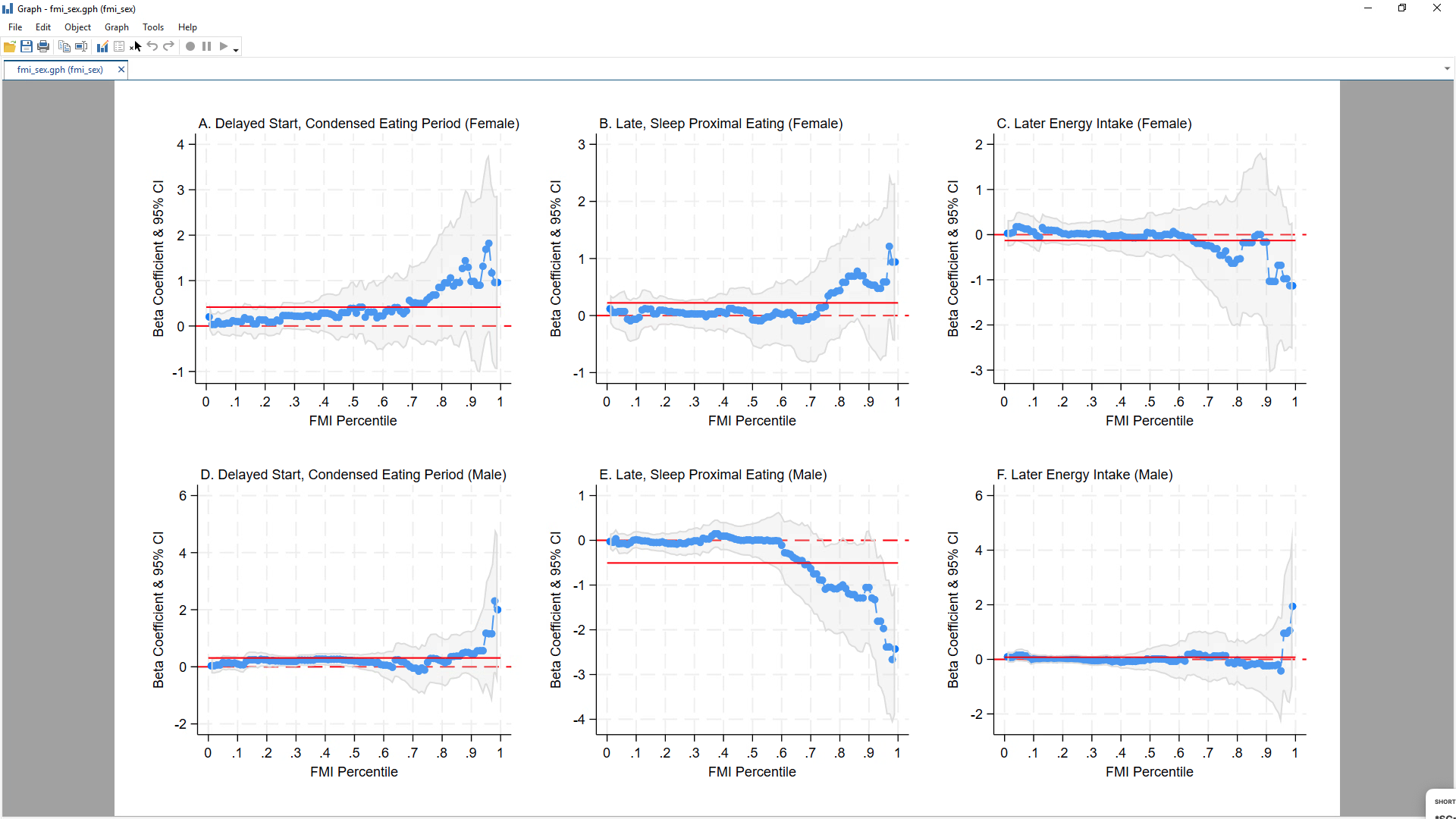

**Supplemental Figure 4.** Quantile regression plots illustrating minimally adjusted associations between three meal timing patterns and FMI by sex. All models were adjusted for age. The red solid line represents the ordinary least squares estimate. The red dotted line indicates the reference line. Abbreviations: FMI, fat mass index.

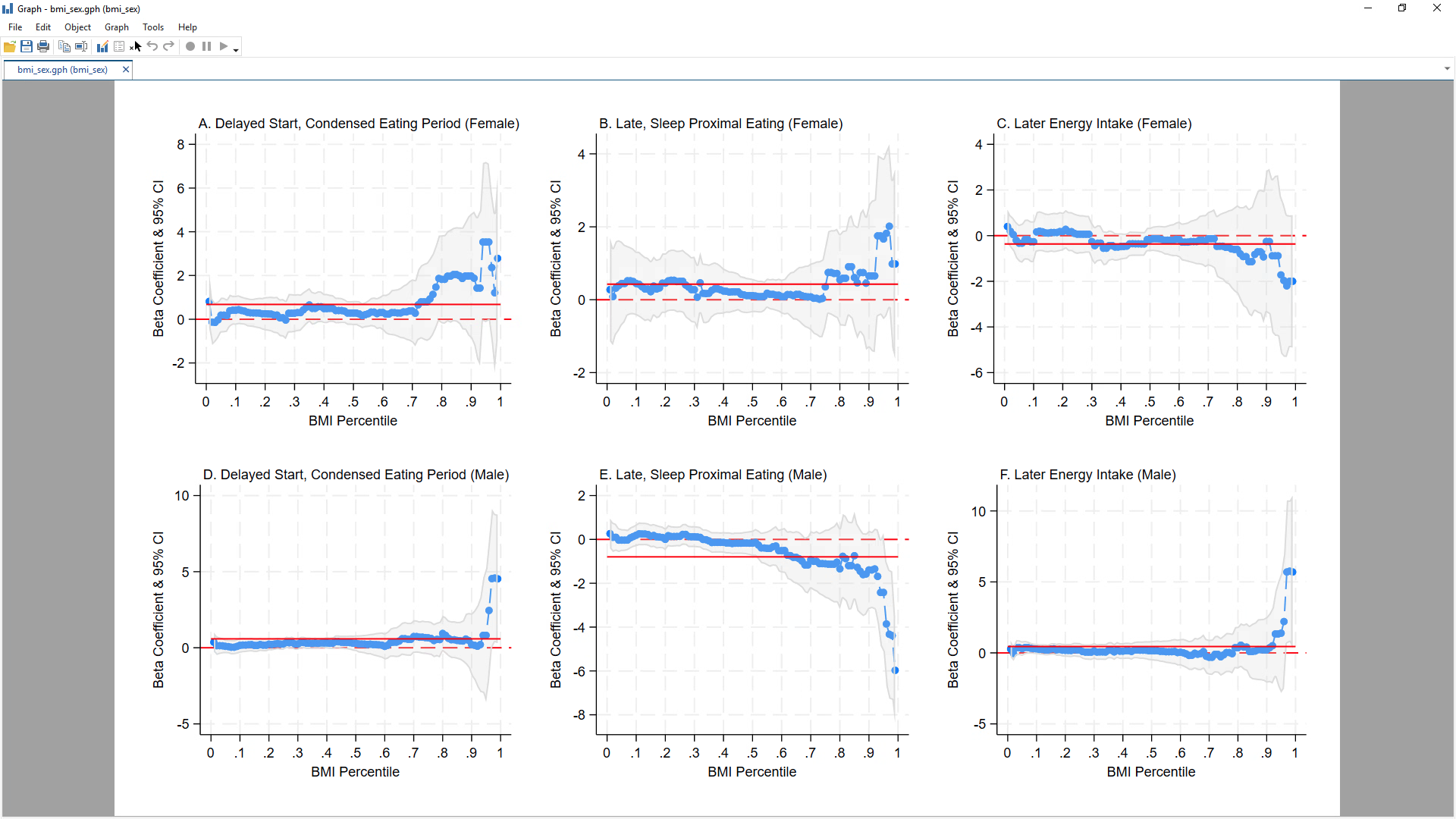

**Supplemental Figure 5.** Quantile regression plots illustrating minimally adjusted associations between three meal timing patterns and BMI by sex. All models were adjusted for age. The red solid line represents the ordinary least squares estimate. The red dotted line indicates the reference line. Abbreviations: BMI, body mass index.

.

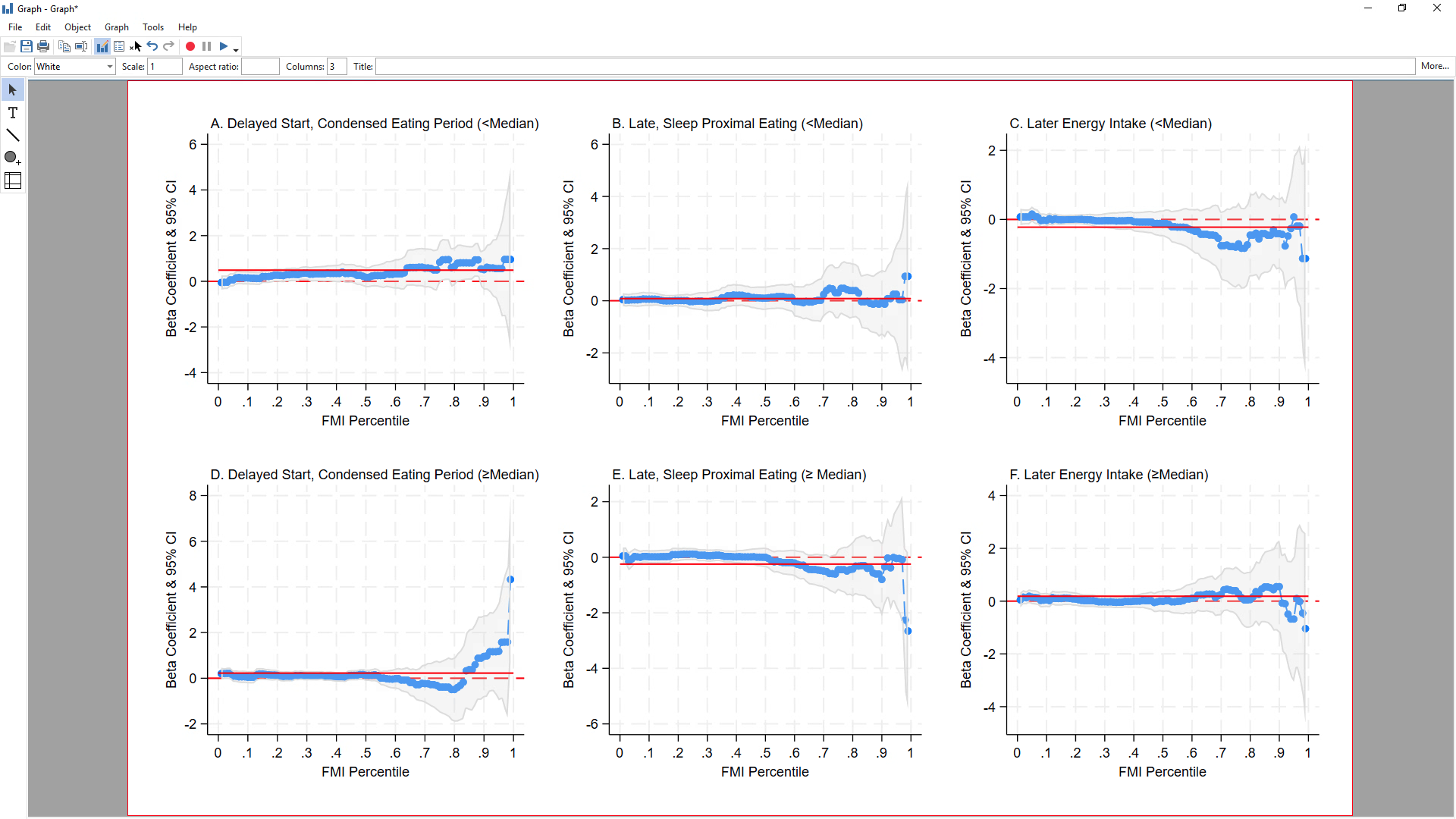

**Supplemental Figure 6.** Quantile regression plots illustrating minimally adjusted associations between three meal timing patterns and FMI by social jet lag categories. All models were adjusted for age and sex. The red solid line represents the ordinary least squares estimate. The red dotted line indicates the reference line. Abbreviations: FMI, fat mass index.

.

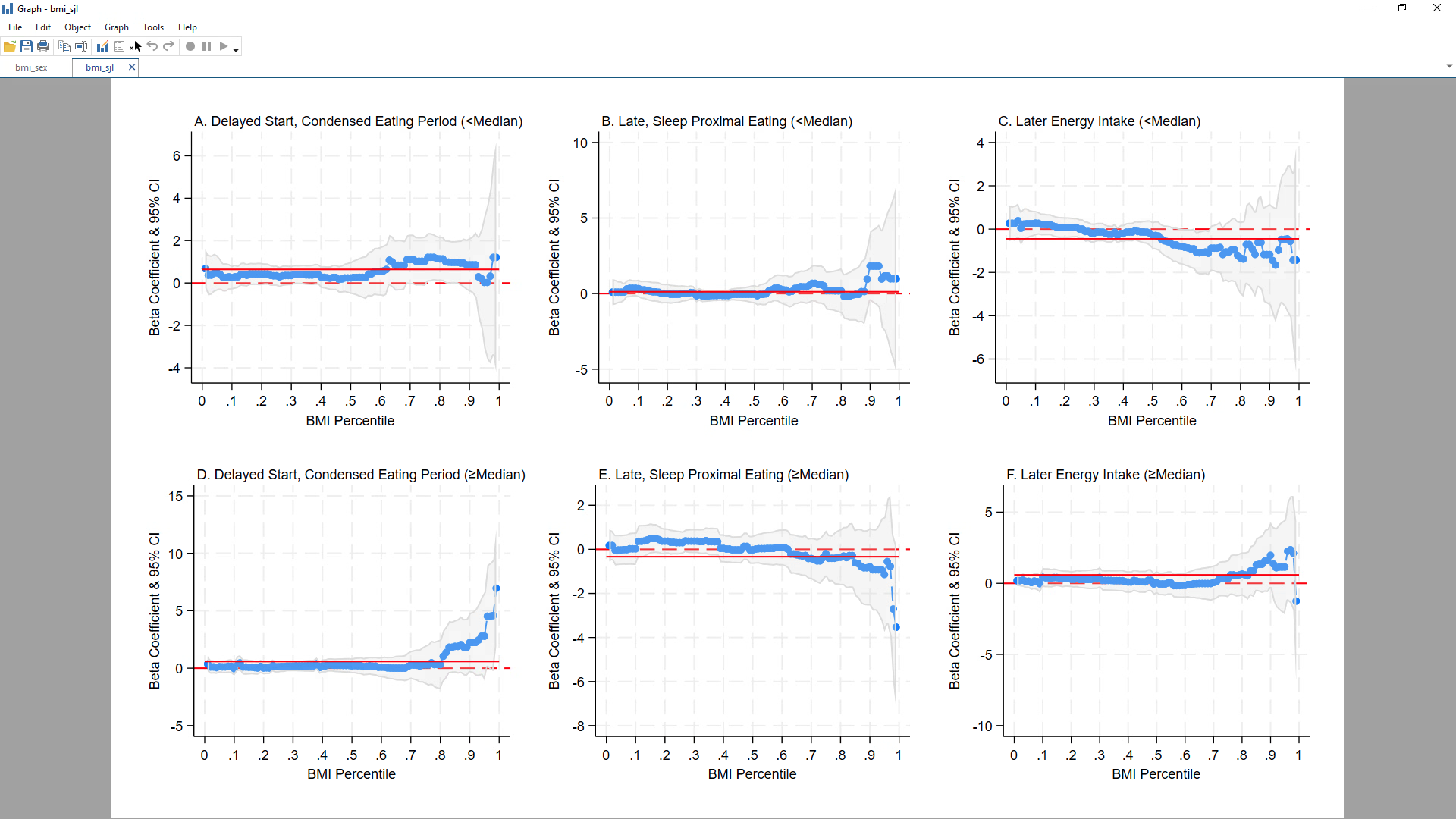

**Supplemental Figure 7.** Quantile regression plots illustrating minimally adjusted associations between three meal timing patterns and BMI by social jet lag categories. All models were adjusted for age and sex. The red solid line represents the ordinary least squares estimate. The red dotted line indicates the reference line. Abbreviations: BMI, body mass index.
